# Non-adherence to surgical antibiotic prophylaxis guidelines: findings from a mixed-methods study in a developing country

**DOI:** 10.1101/2025.04.04.25325227

**Authors:** Noha Ali, Ranim Hamouda, Rana Tarek, Menna Abdelhamid, Abdullah Lashin, Rania Hassan, Rania Gamal, Mourad Elfaham, Aya Attia, Ahmed Abdelaleem, Noha Sakna, Amgad Gamal, Sally Aboelenin, Rahma AbdelHafez, Sara Abdelkader, Ashraf Nabhan

## Abstract

**Objectives:** To assess adherence to surgical antibiotic prophylaxis (SAP) guidelines in obstetric and gynecologic procedures and identify factors contributing to non-adherence.

**Design:** A mixed-methods study comprising a cross-sectional survey and qualitative analysis.

**Setting:** Ain Shams University Hospital of Obstetrics and Gynecology, Egypt.

**Participants:** The survey included data on all surgical procedures performed from November 1, 2024, to December 31, 2024. Eight healthcare providers participated in the qualitative research.

**Methods:** Trained medical interns collected routine data in real-time in the operative theater and in the wards by observing and documenting three key variables namely the antibiotic prescribed, timing of administration, and the duration of use. The overall adherence rate was calculated as the proportion of cases meeting all three criteria. The survey was followed by qualitative research through a synchronous online focus group of eight healthcare providers. Following the transcription of the audio-recorded discussion, three researchers used a deductive approach to content analysis of the focus group discussion.

**Results:** Two handreds and eighty surgical procedures were analyzed, with cesarean sections accounting for 48.6% (136/280). Full adherence to SAP guidelines was observed in 0% of cases. The appropriate antibiotic was prescribed in 62.5% (175/280) of procedures. Timely administration within the recommended 60-minute pre-incision window occurred in 38.2% (107/280). In contrast, 61.4% (172/280) of procedures had delayed antibiotic administration post-incision. The recommended single-dose or 24-hour regimen was administered in only 6.1% (17/280), whereas 93.9% (263/280) had prolonged parenteral antibiotic use beyond 24 hours, with 98.9% (277/280) transitioning to oral antibiotics upon discharge. Key barriers to adherence included knowledge gaps, workflow inefficiencies, inadequate monitoring, limited antibiotic availability, financial constraints, and weak enforcement of SAP guidelines.

**Conclusions:** Non-adherence to SAP guidelines is alarmingly high, particularly regarding timing and duration. Addressing systemic barriers and enforcing guideline compliance is essential to improving antibiotic stewardship in surgical settings.

**Article summary:** *Strengths and limitations of this study:*

- The study investigates adherence to surgical antibiotic prophylaxis guidelines, a critical issue with profound implications for patient safety and antimicrobial stewardship, making it highly valuable for both academic research and clinical practice.
- The study employed a well-structured research design with appropriate data collection techniques, ensuring reliable and valid results. The mixed-methods approach allowed the integration of numerical data with detailed insights from participant experiences, thus providing a more holistic view of the problem. Using multiple data sources enhanced the reliability and credibility of findings by cross-verifying results.
- The study was conducted at a single university hospital in a developing country, which may limit generalizability.
- The work did not assess the risk of postoperative infectious morbidity as the result of non-adherence to surgical antibiotic prophylaxis guidelines.

## Introduction

Surgical antibiotic prophylaxis (SAP) is a key intervention for reducing the risk of postoperative infectious morbidity. Its appropriate use significantly decreases the incidence of surgical site infections (SSIs), which are associated with increased patient morbidity, prolonged hospital stays, and higher healthcare costs. [1,2] SAP effectiveness depends on proper adherence to established guidelines regarding the selection, timing, dosage, and duration of antibiotic administration. [1]

On the other hand, an inappropriate use of SAP contributes to unnecessary costs, adverse events, disruption of normal microbiota, and most critically, to the global burden of antimicrobial resistance (AMR). AMR is responsible for approximately 700,000 deaths annually. [3–6]

Despite clear recommendations, inappropriate use of prophylactic antibiotics continues to be a challenge in many healthcare settings, particularly in low- and middle-income countries (LMICs). [6] The scale of this inappropriate SAP in LMICs remains uncertain due to a lack of accurate and updated data. [7–9]

Egypt’s population ranks number 13 in the list of countries by population. The country’s tertiary hospitals specializing in obstetrics and gynecology perform a high volume of surgical procedures, including cesarean sections and hysterectomies, making adherence to SAP guidelines crucial in minimizing infectious morbidity. Egypt has an up-to-date and clear national guideline for antimicrobial prophylaxis, [10] yet data on adherence to SAP guidelines remain sparse.

This project aimed to assess adherence to SAP guidelines, including antibiotic selection, timing of administration, and duration of use, in an obstetric and gynecology tertiary university hospital. The qualitative part aimed to understand the factors contributing to non-adherence to SAP guidelines in a developing country.

## Methods

The project consisted of two phases: an audit and a focus group discussion. It was prospectively registered on the Open Science Framework (https://osf.io/y7hk9) as a quality improvement initiative. The Institutional Review Board at Ain Shams University reviewed the project and approved it as an exempt protocol. All collected data remained fully deidentified throughout the study.

### The audit

We conducted a cross-sectional survey at the university hospital of the department of Obstetrics and Gynecology, Ain Shams University. We followed the methodology for the Point Prevalence Survey on Antibiotic Use in Hospitals. [11]

Following the design and preparatory phase, data collection started on November 1, 2024 and ended on December 31, 2024. The survey included data on antibiotic prophylaxis of all surgical procedures at the university hospital of obstetrics and gynecology. Exclusion criteria included the need for therapeutic antibiotics. A team of trained interns collected data in real time in the operative theater using a standardized data collection form. Data collection proceeded as a natural task scenario to mitigate the Hawthorne effect. The data collected did not contain any identifiable information, and there is no way to link the information back to identifiable information. The team stored data in an online university database created specifically for this project. The design of the form was by the Point Prevalence Survey on Antibiotic Use in Hospitals [11] and included the following variables: surgical details (type of surgery and its urgency), and antibiotic prophylaxis parameters (antibiotic choice, timing of administration, dosage, duration, antibiotic count, and route of administration).

Adherence to SAP guidelines was assessed using three parameters: antibiotic selection, timing of administration, and duration of prophylaxis. Appropriateness of antibiotic choice was based on hospital or national guidelines. Timing of administration was defined as optimal if given within 60 minutes before incision. Duration was deemed appropriate if limited to a single preoperative dose or 24 hours post-operative. The overall adherence rate was calculated as the proportion of cases meeting all three criteria.

The adherence rate was presented as a proportion. Descriptive data analysis was performed using R v4.4. [12]

### The qualitative study

We invited eight healthcare providers to participate in a synchronous online focus group (SOFG). The members of the group were of different age groups, specialties, and cadres. We maintained gender balance in the group. All participants signed informed consent to participate in the study, to the use of their data and opinions in the focus groups, as well as to their audio-recording, and were informed that they can withdraw at any time.

We used the MS Teams application to organize the SOFG on March 25, 2025. Eight healthcare providers participated in the focus group discussion (FGD). The duration of the meeting was 60 minutes. The lead researcher (male, professor) moderated the FGD, and the other (female, research fellow) took notes and asked follow-up questions. At the start of the FGD, the moderator explained the purpose of the meeting and then introduced the open-ended question: What are the factors contributing to SAP non-adherence? Members of the focus group provided their insights. All participants had a chance to share, and the recording included the input from each participant. The moderator promoted the discussion and maintained neutrality while ensuring an interactive atmosphere and contributions from every participant.

The FGD was transcribed verbatim and examined by three researchers (SAA, SIA, and RAA) who had no prior acquaintance with the focus group members. We used a deductive approach to content analysis of the FGD. This approach involved systematically analyzing textual data based on predefined themes. [13] The predefined themes included health workforce, service delivery, health information systems, medical products, financing, and leadership and governance.

The three researchers initially reviewed the transcripts thoroughly before extracting and coding meaning units. The codes were then grouped into the pre-specified themes. Each step was completed individually before the researchers collaboratively reviewed the analysis and reached a consensus. While the primary focus was on fitting data into existing categories, the three researchers remained open to minor modifications if new insights emerged. [14–17]

Following the completion of the content analysis, the findings from the focus group discussion were triangulated with the survey findings to explore any convergence. [18,19]

### Public involvement

Members of the public were not involved directly in the design of this project. Although not explicitly part of this project, the main idea of this work was definitely inspired by the daily discussion between doctors and members of the public regarding the burden of misuse of antibiotics.

## Results

Data on 280 obstetric or gynecologic surgical procedures were collected. Residents of obstetrics and gynecology and anesthesia were responsible for the preparation of the operative procedures, including the provision of antibiotic prophylaxis. Almost half of the procedures were cesarean sections (136/280 [48.6%]), followed by hysterectomies (50/280 [17.9%]). Elective procedures accounted for 93.9% (263/280) of the work, Table 1.

**Table 1:**
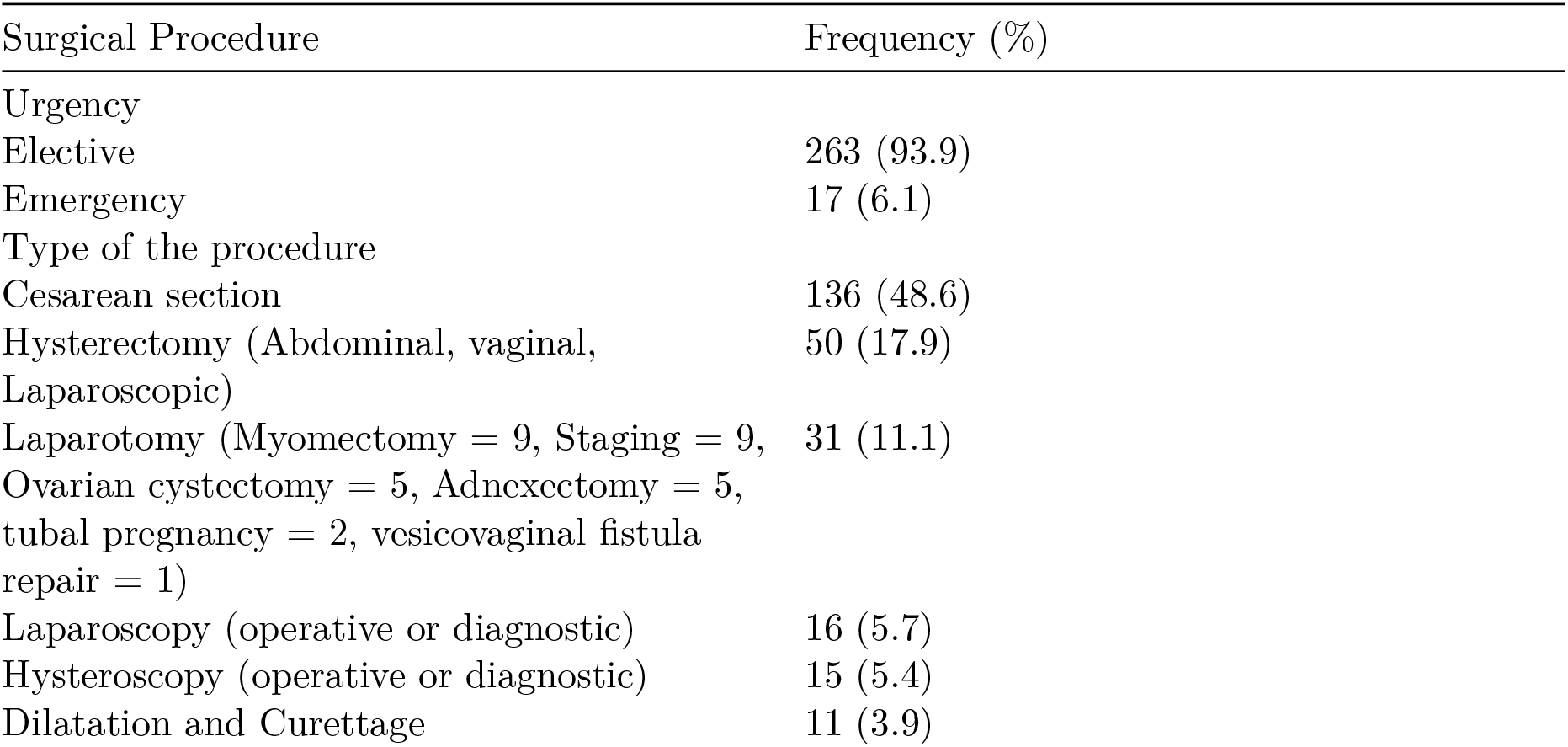
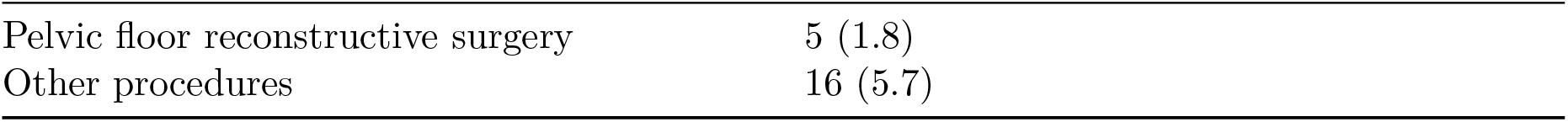
Surgical Procedures.

### Adherence to Surgical Antibiotic Prophylaxis Guidelines

Intravenous antibiotic prophylaxis was administered in all procedures. Continuous intravenous infusion was used in 66.1% (185/280) and intermittent intravenous injections in 33.9% (95/280) of procedures.

Non-indicated SAP was observed in 8.2% (23/280) of cases. This included eleven dilatation and curettage, five diagnostic Hysteroscopies, three diagnostic laparoscopies, two hymenotomies, one cervical cerclage, and one examination under anesthesia.

### Overall adherence

Overall adherence to all SAP guidelines - correct antibiotic selection, timing, and duration - was not observed in any of the procedures.

### Antibiotic Selection

The appropriate antibiotic, as per hospital or national guidelines, was administered in 62.5% (175/280) of procedures. In the remaining 37.5%, either a non-recommended antibiotic or a combination of antibiotics was administered. All deviations were due to the absence of the recommended antibiotic at the time of procedure.

A single antibiotic was used in 65.7% of procedures (184/280). Multiple antibiotics were used in 34.3% of procedures (96/280). A cephalosporin was administered in most procedures (255/280 [91.1%]), either as a single agent or in combined regimens. Metronidazole was the most frequently added antibiotic (41 out of the 96 combined regimens), Table 2.

**Table 2:** Antibiotic Selection.

| Antibiotic Choice | Frequency |
| --- | --- |
| **Single agent** | 184 |
| Cefoxitin | 112 |
| Cefazolin | 46 |
| Ampicillin-Sulbactam | 17 |
| Cefotaxime | 4 |
| Meropenem | 2 |
| Cefoperazone-Sulbactam | 1 |
| Ceftriaxone | 1 |
| Metronidazole | 1 |
| **Multiple agents** | **96** |
| Cefoxitin, Cefazolin | 24 |
| Cefoxitin, Metronidazole | 14 |
| Cefoxitin, Ampicillin-Sulbactam | 14 |
| Cefoxitin, Cefazolin, Metronidazole | 10 |
| Cefazolin, Ampicillin-Sulbactam | 5 |
| Cefazolin, Metronidazole | 5 |
| Cefoxitin, Cefotaxime | 5 |
| Ampicillin-Sulbactam, Metronidazole | 4 |
| Cefotaxime, Cefazolin | 3 |
| Cefotaxime, Clindamycin | 2 |
| Cefoxitin, Ampicillin-Sulbactam, Metronidazole | 2 |
| Cefazolin, Ampicillin-Sulbactam, Metronidazole | 1 |
| Cefazolin, Ampicillin-Sulbactam, Clindamycin | 1 |
| Cefazolin, Cefoxitin, Metronidazole | 1 |
| Cefotaxime, Ampicillin-Sulbactam, Metronidazole | 1 |
| Cefotaxime, Metronidazole | 1 |
| Cefoxitin, Azithromycin | 1 |
| Cefoxitin, Cefotaxime, Metronidazole | 1 |
| Meropenem, Clindamycin | 1 |

### Timing of Administration

Antibiotics were administered within the recommended 60-minute window before incision in 38.2% (107/280) of procedures. In contrast, 61.8% of cases experienced delayed or premature antibiotic administration.

The first dose was administered after the start of surgery in 61.4% (172/280) of procedures. The inappropriate timing of the first dose was not significantly more common in emergency (75%) compared to elective procedures (60.8%) (p = 0.3).

### Duration of Prophylaxis

The recommended single-dose or 24-hour regimen was adhered to in only 6.1% (17/280) of procedures. Prolonged parenteral antibiotic use in hospital beyond 24 hours was observed in 93.9% (263/280) of procedures. A switch to oral antibiotics on hospital discharge was observed in 98.9% (277/280) of cases. Outpatient oral antibiotics continued for up to seven days following discharge.

### Factors Contributing to Non-Adherence to SAP Guidelines

A deductive approach to qualitative content analysis was performed by applying predefined themes (health system building blocks) to examine factors influencing SAP non-adherence.

#### Theme 1: Health Workforce

Workforce-related factors include inadequate training, knowledge gaps, and limited accountability regarding SAP guidelines. Participants reported inconsistent adherence due to a lack of awareness or confusion about best practices.

These quotes illustrate how workforce-related factors contribute to non-adherence.

“Not everyone knows the exact timing and dose of prophylactic antibiotics. Some just go by what they’ve always done.”

“New staff don’t receive structured training on SAP; they learn on the job, which leads to inconsistencies.”

“There’s a real lack of knowledge and proper monitoring when it comes to using antibiotics.”

“Non-adherence is more about not knowing and not following the existing guidelines than it is about the guidelines themselves being missing.”

“Junior residents don’t get any formal training on surgical antibiotic prophylaxis; they learn with hands-on experience, which leads to inconsistencies.”

“Some healthcare providers aren’t clear on the appropriate timing and duration for prophylactic antibiotics, often sticking to their usual habits instead of following standardized protocols.”

#### Theme 2: Service Delivery

Service delivery is the way in which healthcare is being provided, including readiness to provide SAP in a timely fashion. Service delivery factors impacting SAP adherence include workflow inefficiencies, time constraints, and lack of standardized protocols. Participants described how the inefficient process of dispensing prophylactic antibiotics, lack of coordination among teams, and high patient volumes created challenges in adhering to SAP guidelines.

The following quotes highlight the structural and procedural challenges in service delivery that hinder appropriate SAP administration.

“There’s no clear protocol followed by all teams, so practices vary from one department to another.”

“We are understaffed, and in the rush to prepare for surgery, SAP timing is often overlooked.”

“The high patient flow in our hospital makes it difficult to maintain adherence to the guidelines for surgical antibiotic prophylaxis.”

“Surgical antibiotic prophylaxis tends to be viewed as one of the less critical elements of patient care in the operative theater, with other surgical priorities taking the spotlight. This mindset can result in insufficient focus on its administration, even though it plays a vital role in preventing infections after surgery.”

“While junior doctors may be aware of the guidelines, they often overlook their importance and follow the outdated practices from their seniors, even when they’re wrong.”

“in the operative room, we do not follow a mandatory checklist for the timely pre-incision administration of SAP.“

#### Theme 3: Health Information Systems

Information is essential for monitoring and evaluation. Inadequate documentation and lack of real-time monitoring systems hinder adherence to SAP guidelines. Participants noted that inconsistent record-keeping prevents effective tracking and evaluation of SAP compliance.

The following quotes demonstrate challenges related to health information systems.

“SAP administration is not always documented properly, so we don’t have reliable data to track compliance.”

“There’s no alert system to remind staff when to administer prophylactic antibiotics.”

“Records are sometimes incomplete, making it difficult to assess whether SAP guidelines were followed.”

“There’s no system in place to check whether antibiotics were administered correctly.”

#### Theme 4: Medical Products

This refers to equitable access to essential pharmaceutical products of assured quality, safety, efficacy, and cost-effectiveness and their scientifically sound and cost-effective use. Limited availability of appropriate antibiotics and stock-outs negatively impact adherence. Participants described instances where they had to use alternative antibiotics due to supply chain issues.

These quotes reflect how access to medical products affects adherence to SAP guidelines. “Sometimes, the recommended antibiotic is unavailable, so we use whatever is in stock.”

“Non-adherence to prophylactic antibiotics is more often due to a lack of the recommended antibiotics and accepted alternatives rather than a lack of knowledge.”

“The occurrence of adverse events and the concerns regarding the quality of the available antibiotics and poor storage complicate things even more and drive doctors to use a trusted antibiotic even when it is not the one recommended by guidelines.”

#### Theme 5: Financing

Financial constraints limit SAP adherence by affecting resource availability, staff training, and procurement of necessary medical supplies. Participants noted that budget limitations reduce access to quality antibiotics and delay system improvements.

The following quotes illustrate how financial barriers influence SAP adherence.

“Budget cuts mean we can’t always afford the recommended antibiotics, so we use cheaper alternatives.”

“There’s no dedicated funding for continuous training on SAP guidelines.”

“Financial constraints affect every aspect, from procurement to training to electronic systems, making it difficult to implement SAP guidelines correctly.”

“The model of unified governmental procurement sometimes means that we do not get the recommended antibiotics, so we might be compelled to use whatever they send.”

#### Theme 6: Leadership and Governance

This involves ensuring that policy frameworks exist and are combined with effective oversight, regulation, attention to system design, and accountability. Weak governance structures and lack of enforcement of SAP guidelines contribute to non-adherence. Participants expressed concerns about the absence of accountability mechanisms and inconsistent policy implementation.

The following quotes highlight governance-related barriers to SAP adherence. “There’s no strict enforcement of SAP guidelines, so practices vary widely.”

“Leadership hasn’t prioritized SAP compliance, so there’s little motivation to follow protocols strictly.”

“Policies exist on paper, but in practice, adherence is not closely monitored or evaluated.”

“Having a national guideline is only a starting point; hospitals need to have their own clinical protocols based on reliable, trusted evidence rather than personal opinions. For instance, one hospital’s protocol once stated that IUD insertion required antibiotic administration, despite compelling evidence to the contrary.”

“The anxiety surrounding potential legal consequences often pushes doctors into practicing defensive medicine. Many healthcare providers tend to over-prescribe antibiotics or ignore the established guidelines due to the fear of legal repercussions, which can lead to unnecessary use and issues with surgical antibiotic prophylaxis.”

“The culture of blame compels many junior doctors to extend postoperative antibiotic use. If a patient develops a surgical site infection, we are blamed—even when we strictly follow SAP guidelines. However, overprescribing antibiotics rarely attracts criticism, making it the safer choice in a blame-driven environment.”

### Triangulation of quantitative and qualitative findings

Overall, the non-adherence can be explained by knowledge gaps, workflow inefficiencies, lack of real-time monitoring systems, limited availability of appropriate antibiotics, financial constraints, and lack of enforcement of SAP guidelines, Table 3

**Table 3:** Triangulation of quantitative and qualitative findings.

| Theme | Qualitative Findings | Survey Data<br>(Quantitative) | Triangulation |
| --- | --- | --- | --- |
| Workforce | Lack of awareness and confusion about best practices | No adherence to all three parameters | Convergent |
| Service Delivery | Workflow inefficiencies and high patient volume | Delayed administration in 61.4% of cases | Convergent |
| Health Information Systems | Poor documentation and lack of real-time monitoring | No adherence to all three parameters | Convergent |
| Medical Products | Stock shortages of recommended antibiotics | 37.5% received non-recommended antibiotics | Convergent |
| Financing | Limited resources for training and procurement | 93.9% prolonged use beyond 24 hours | Convergent |
| Governance | Weak enforcement and non-compliance culture | Outpatient oral antibiotics prescribed in 98.9% of cases | Convergent |

### Proposed Actions to Overcome Barriers

The focus group identified several key actions to address barriers in our setting:

1. Enhancing the consistent implementation of SAP: Participants suggested the following ideas to improve access to evidence-based recommendations:
  - Developing and disseminating standardized SAP hospital protocols that are aligned with the national guidelines.
  - A printed or electronic copy of the SAP protocol should be made available to all healthcare providers in the hospital.
2. Addressing Educational Gaps: Participants highlighted the critical role of knowledge translation and utilization by conducting regular training and orientation sessions. Training should target surgeons, anesthetists, nurses, and clinical pharmacists and should cover all aspects of appropriate antibiotic selection, timing of administration, and duration of SAP.
3. Optimizing Workflow Efficiency: Participants emphasized the need to
  - Integrate SAP into a preoperative checklist.
  - Have a stock of the recommended antibiotic available in operating rooms to prevent delayed administration.
  - Establish a reminder for SAP in the operating rooms to ensure timely administration.
4. Strengthening Multidisciplinary Collaboration: Participants highlighted the need to establish a functional antimicrobial stewardship team to actively monitor compliance and improve SAP adherence.
5. Implementing Audit and Feedback Mechanisms: Participants emphasized the need to
  - Conduct regular audits and present compliance data at surgical team meetings.
  - Provide real-time feedback to clinicians to reinforce best practices.
6. Promoting Accountability and a Supportive Culture: Participants highlighted the need to cultivate a culture where all team members, from nurses to pharmacists, feel empowered to question non-compliant practices.

## Discussion

This mixed-methods study assessed adherence to surgical antibiotic prophylaxis guidelines in one of the largest tertiary university hospitals of obstetrics and gynecology in Egypt. We sought to identify gaps in practice and potential areas for improvement. The findings revealed that while the recommended antibiotic selection was usually implemented, substantial deviations from standard guidelines were observed in the timing and duration of antibiotic administration. Adherence to all three SAP parameters was never attained in any of the included procedures. Key barriers to adherence included knowledge gaps, workflow inefficiencies, inadequate monitoring, limited antibiotic availability, financial constraints, and weak enforcement of SAP guidelines.

Our comprehensive literature search clearly showed the paucity of data from Egypt. Our data showed that the most common deviation from evidence-based recommendations was the prolonged antibiotic use beyond 24 hours. In almost all procedures, we observed a switch to oral antibiotics for up to seven days after discharge from hospital. The current status is probably worse than previously reported one decade ago. [20,21] This overuse of antibiotics in developing countries has been recently documented in a rigorous scoping review. [22] Extending antibiotic use beyond 24 hours does not reduce the risk of postoperative infectious morbidity but increases the likelihood of antimicrobial resistance and adverse events [5]. The fear of infections, fear of litigation, misconceptions regarding antibiotic prophylaxis, the culture of blame, and the practice of defensive medicine contributed to this disturbing practice.

Timely administration of prophylactic antibiotics within the recommended 60-minute window before surgical incision is crucial for optimal efficacy. [5,7] In our survey, only 38.2% of patients received antibiotics at the appropriate time. The limited literature from developing countries on adherence to this 60-minute window suggests the same poor practice. [20,21] Administration timing (0–60 minutes before incision) is the most problematic indicator even in developed countries. [23] We observed lapses in timing as a result of workflow inefficiencies, anesthesia-related delays, and inadequate awareness among healthcare providers.

Studies have shown variability in compliance with SAP protocols, influenced by factors such as institutional policies, healthcare provider knowledge, and resource availability. [24] An audit of antibiotic prophylaxis in the Eastern Mediterranean Region revealed that many hospitals lack standardized SAP protocols or fail to enforce existing ones. The same audit indicated that surgeons and other healthcare providers may not always be fully aware of or adhere to updated SAP guidelines, leading to inconsistent practices. [25]

A recent qualitative study from a high-income country demonstrated that a lack of structured workflow was the main factor contributing to non-adherence to antibiotic guidelines in the peri-operative settings. [26] We believe that in LMICs, the main factors have a different profile. Our study indicates that weak enforcement of the guidelines, occasional unavailability of recommended antibiotics, lack of audits or regular feedback, and a lack of fidelity for implementing the guidelines were the main barriers to adherence to SAP guidelines. Some clinicians resist adopting guidelines, especially if previous practices were perceived as effective. Providers expressed their belief that guidelines do not fit the nature of a public hospital serving poor patients with inadequate self-hygiene and poor living conditions at home. Providers extended antibiotic duration beyond guidelines as a precautionary measure, despite their knowledge of the evidence against it. Fear of infections and litigation and consequently the practice of defensive medicine probably contributed to the overuse of antibiotics as a safety measure.

While the overall compliance with guidelines is around 64% in developed countries [23], the picture is different in developing countries, as shown by our data. We understand the numerous challenges for implementing effective and sustainable antibiotic stewardship programs in developing countries. [27] The implementation of effective and sustainable antibiotic stewardship programs in developing countries is not impossible. A recent systematic review assessed the potential for a successful implementation of such programs in African countries, including Egypt. The successes reported in some countries may give us some hope that other developing countries can implement these programs. [28]

## Limitations

The findings of this work should be interpreted in the context of its limitations. The survey was conducted at a single university hospital, which may limit generalizability. This work did not assess the risk of postoperative infectious morbidity among non-adherent participants. Future studies should include multicenter data. Future studies should assess the impact of targeted interventions, such as provider education, regular feedback, and electronic alerts, on SAP adherence in low-resource, high-flow settings.

## Conclusion

The lack of overall adherence to key parameters of antibiotic prophylaxis in one of the largest developing countries is alarming. Efforts should focus on the appropriate timing and duration of administration. This calls for a sustained program with strict enforcement.

## Data Availability

All data relevant to this project are publicly available. The data, analysis script and materials related to this project are publicly available in the project folder in OSF.

https://osf.io/y5bgu

## Declarations

### Ethics approval

This Quality Improvement project was reviewed by the Institutional Review Board at Ain Shams University and was approved an exempt protocol. All data remained deidentified.

### Consent for publication

Not applicable.

## Acknowledgments

We sincerely thank the healthcare providers at Ain Shams University Hospital of Obstetrics and Gynecology for their dedication and commitment to patient care in the operating rooms. We deeply appreciate their contributions and the valuable role they play in ensuring high-quality surgical care.

## Author contributions

CRediT authorship contribution statement: **Noha Ali**: Writing - review & editing, Project administration, investigation. **Ranim Hamouda**: Writing - review & editing, Investigation. **Rana Tarek**: Writing - review & editing, investigation. **Abdullah Lashin**: Investigation. **Rania Hassan**: Writing - review & editing, Investigation, Supervision. **Rania Gamal**: Investigation, Supervision. **Menna Abdelhamid**: Investigation. **Mourad Elfaham**: Writing - review & editing, Investigation, Supervision. **Aya Attia**: Writing - review & editing, Supervision. **Ahmed Abdelaleem**: Writing - review & editing, Investigation, Supervision. **Noha Sakna**: Writing - review & editing, Supervision. **Amgad Gamal**: Investigation, Supervision. **Sally Aboelenin**: Writing - review & editing, Investigation, Data analysis. **Rahma Abdel- Hafez**: Writing - review & editing, Investigation, Data analysis. **Sara Abdelkader**: Writing - review & editing, Investigation, Data analysis. **Ashraf Nabhan**: Writing - original draft, Methodology, Supervision, Data analysis, Conceptualization.

